# Pharmaco-psychiatry and gut microbiome: A systematic review of effects of psychotropic drugs for bipolar disorder

**DOI:** 10.1101/2024.12.09.24318729

**Authors:** Truong An Bui, Benjamin R. O’Croinin, Liz Dennett, Ian R. Winship, Andrew Greenshaw

## Abstract

Despite being one of the most common and debilitating mood disorders, bipolar disorder is often misdiagnosed and undertreated. Its pathogenesis is complex, with significant patient variability and inconsistent treatment effectiveness. The brain-gut-microbiota axis plays a critical role in bipolar disorder by modulating neurotransmitter secretion, gut peptides, and systemic inflammation. However, the mechanisms by which psychotropic treatments influence gut microbiota composition and their implications for clinical outcomes remain poorly understood. This systematic review evaluated the impact of psychotropic drugs on gut microbiota and their potential role in bipolar disorder treatment outcomes.

A comprehensive search across Ovid MEDLINE, Embase, APA PsycINFO, Scopus, and PubMed yielded 314 articles, of which 12 met the inclusion criteria (last search: 13 August 2024). The studies included were those on adults with bipolar disorder type I or II receiving psychopharmacological treatments, those with group comparisons (e.g., medicated vs. non-medicated) investigating gut microbiome changes; no restrictions applied to psychotic features, comorbid anxiety, or prior treatment responses. Exclusions involved individual case reports, incomplete conference submissions, or early terminated studies lacking efficacy analysis. Cochrane ROBINS-I V2 tool was used to measure the risk of bias, and the GRADE approach was utilized to rate the certainty of evidence in included studies. Two authors independently extracted data into Excel spreadsheets, categorizing demographic and clinical characteristics, describing microbiome analytic methods, and summarizing findings on gut microbiome changes post-treatment. Given the high variability in methods and outcome measures across studies, all details were reported without data conversion.

Data synthesis reveals that psychotropic treatments, including quetiapine and lithium, influence gut microbiota by increasing the abundance of beneficial bacteria supporting gut health and pathogenic bacteria linked to metabolic dysfunction. Notably, female patients exhibited more significant changes in microbial diversity following psychotropic treatment. Additionally, patients treated with psychotropics showed an increased prevalence of gut bacteria associated with multidrug antibiotic resistance. In bipolar patients treated with quetiapine, responders—those experiencing improved depressive symptom scores—displayed distinct gut microbiome profiles more closely resembling those of healthy individuals compared to non-responders. Responders also exhibited neural connectivity patterns similar to healthy subjects. These findings underscore the complex dual impact of psychotropic medications on gut microbiota, with potential consequences for both gut and mental health. While the enrichment of beneficial bacteria may support gut health, the rise in antibiotic-resistant and metabolically disruptive bacteria is concerning.

Study limitations include methodological heterogeneity, a high risk of bias in some studies due to incomplete statistical analyses or insufficient control for confounding factors, and potential duplication of study populations arising from overlapping authorship. Further research is essential to elucidate the functional consequences of these microbial shifts and their influence on treatment efficacy. Nevertheless, this review highlights the potential of utilizing gut microbiota profiles to inform personalized treatment strategies, optimize therapeutic outcomes, and minimize side effects in bipolar disorder.

## Introduction

### Gut-brain axis crosstalk and mental illness: an overview

Current interest in a holistic view of nervous system function, including internal and external environmental modulators, now includes the bidirectional crosstalk between the gut and the brain, which is now referred to as the gut-brain axis. This review focuses on the relevance of the gut-brain axis to understanding mechanisms underlying mental illness, with a specific focus on the effects of drugs used to treat bipolar disorder (BP), a prominent and disabling mood disorder.

The gastrointestinal tract contains the largest, most abundant and diverse microbial reservoir in the human body, which serves as the first line of immune defence (1–3). Gut microbiota comprises four major bacteria: *Bacteroidetes*, *Firmicutes*, *Proteobacteria*, and *Actinobacteria* (4–6). This microbiome can modulate the hypothalamic-pituitary-adrenal (HPA) axis, which mediates neuroendocrine and autonomic functions (7,8). With digestion, the gut microbiome produces an abundance of neuroactive metabolites such as neurotransmitters and their precursors, including glutamate, noradrenaline, serotonin (5-HT), dopamine (DA), and gamma-aminobutyric acid (GABA), tyramine, phenylethylamine, and tryptamine that can regulate brain functions and behaviour (9,10). These neurotransmitters play crucial roles in neurological and immunological activities in the brain (11). Disturbed intestinal microbiota, often associated with reduced bacterial diversity, is associated with a variety of psychiatric diseases, including major depressive disorder (MDD) (12–14), BP (15), and schizophrenia (SCZ) (16). Gut microbiome composition is especially sensitive to stressors in the central nervous system (CNS) that can activate the sympathetic nervous system and HPA axis, such as aging, infection, and injuries (17,18). Stress and depression are known to increase gut barrier permeability and cause epithelial barrier defects, or “leaky gut,” which results in gut bacteria, such as *E. coli* and their lipopolysaccharide, translocating into circulatory fluids within the body and evoking an inflammatory response (15,19,20). These can further promote the occurrence and progression of psychiatric disorders (21,22). However, mechanisms for these relationships have yet to be identified (23)

### Bipolar disorder epidemiology and pathophysiology

BP is the 17th leading contributor to the global burden of disease, just after MDD, anxiety disorders, SCZ, and dysthymia, affecting >1% of the worldwide population (24,25). In 2019, BP was reported to account for 8.5 million global disability-adjusted life-years (DALYs) (26). The global economic burdens and healthcare costs of BP are enormous, as they affect not only the patients but also family members and caregivers (27–29).

BP is characterized by biphasic mood episodes of mania (or hypomania) and depression, which result in extreme and unpredictable emotion, thought, and energy disturbances (30,31). Mania can manifest as hyperactivity, irritability, grandiosity, decreased need for sleep, increased talkativeness, distractibility, delusions, flight of ideas, psychomotor agitation, risk-taking behaviour, and highly elevated mood (32,33). In contrast, depressive episodes are characterized by sadness, reduced interest or pleasure, fatigue, social withdrawal, low self-esteem, psychomotor retardation or agitation, sadness, insomnia or hypersomnia, and change in appetite and weight (33,34). Diagnoses of BP in the Diagnostic and Statistical Manual of Mental Disorders (DSM-V-TR) include bipolar I disorder (BP-I), bipolar II disorder (BP-II), cyclothymic disorder, and substance/medication-induced BP (35).

Medications are generally the most critical component of BP treatment (36,37). Pharmacological therapies used for BP can be divided into mood stabilizers, antipsychotics, and anticonvulsants (also known as antiepileptics) (38,39). During acute treatment, a mood state (e.g., depression) may transition or spill over to its opposite (e.g., mania), adding complexity to clinical decisions regarding psychotropic selection and dosing (40–42). This mood switch process is a distinctive feature of BP compared to other psychiatric conditions, presenting a unique challenge in its management (43). Additionally, among patients receiving pharmacotherapy, up to 35% relapse within the first year in adults, and the risk of relapse rises to 65% after 4-5 years (36,37,44,45).

Functional imaging, such as functional magnetic resonance imaging (fMRI) and positron emission tomography (PET), offer a powerful means of investigating the interplay between the gut microbiome and brain activity in BP. Imaging can identify biomarkers that differentiate BP from other psychiatric conditions, such as unipolar depression or SCZ, which often present overlapping symptoms (46). It also allows researchers to assess the impact of pharmacological treatments (e.g., mood stabilizers) and psychotherapy on brain function, providing insights into therapeutic mechanisms, predicting treatment outcomes response, and creating tailored interventions (47–49).

Techniques like PET enable the study of neurotransmitter systems, such as DA and 5-HT, which are implicated in the pathophysiology of BP (50–52). fMRI can identify abnormal connectivity in brain networks like the default mode network (DMN), prefrontal cortex (PFC), hippocampus, and amygdala. These networks govern self-referential thinking and emotional processing and are disrupted in BP (53–56). For example, compared to healthy controls (HC), BP patients have greater functional connectivity (FC) between the salience and DMN (57) and altered activation within the DMN (53,55,58).

5-HT is a pivotal neurotransmitter in mood stabilization, and its dysregulation plays a significant role in the manic and depressive symptoms of BP (59–61). Dysregulation of 5-HT metabolism has been associated with intestinal microbiota-induced inflammation (62–65) and these neurotransmitter abnormalities can be detected through functional imaging techniques such as fMRI and PET (66–68). Previous studies showed that gut dysbiosis can divert tryptophan, the precursor for 5-HT synthesis, into the kynurenine pathway, thereby reducing 5-HT availability while increasing neurotoxic metabolites (69–71). Additionally, short-chain fatty acids (SCFAs) produced by gut bacteria can affect the serotonin transporter expression and 5-HT receptor sensitivity, which regulates 5-HT reuptake (72). Functional imaging can track changes in glucose metabolism in brain regions affected by gut-derived metabolites, offering a direct visualization of how gut health shapes brain energy utilization (73,74).

Psychotropic agents used to treat BP patients are known to induce weight gain and other metabolic disorders such as obesity, insulin resistance, dyslipidemia, hypertension, impaired glucose tolerance, and high rates of atherosclerotic disease (75–84). These side effects suggest a connection between BP pathophysiology/treatment and metabolism, as well as the gut microbiome. Thus, a comprehensive understanding of the interactions between the gut microbiome, pharmacotherapy, and treatment outcomes is vital. Although the relationship between BP and the gut microbiome is still relatively unexplored, available evidence describing differences in gut microbiome composition between BP and HC has been summarized fairly recently (85–88). To our knowledge, this is the first systematic review of the potential effects of psychotropics on the microbiome of treated and untreated BP individuals.

### Objectives

As the first systematic review on the topic, this review investigates how pharmacological treatments might modulate inflammatory processes, metabolic functions, and neurotransmitter signalling pathways within the gut-brain axis in BP. Gut microbiome in psychiatric disorders is a very complex and rapidly evolving field, and BP is a heterogeneous disorder, with individuals often experiencing varying responses to pharmacological treatments. BP patients also frequently develop comorbidities, including metabolic syndrome, gastrointestinal issues, and other psychiatric conditions. Exploring the role of the gut-brain axis in the context of BP pharmacotherapy provides a novel lens through which these discrepancies and complexities might be explained and lead to more holistic approaches to managing BP. Moreover, understanding these connections could also pave the way for novel personalized therapeutic strategies, inspiring further studies and clinical trials focused on microbiome-targeted interventions.

## Material and Methods

### Inclusion criteria

This review followed the Preferred Reporting Items for Systematic Reviews and Meta-Analyses (PRISMA) guidelines, and the PRISMA 2020 checklist was used (89). The PRISMA 2020 checklist and abstract checklist are available in **S3** and **S4 Table**. The inclusion criteria for studies were as follows: (1) studies of adults diagnosed with BP-I or BP-II; (2) participants received psychopharmacological BP intervention (e.g., mood stabilizers, antipsychotics, antidepressants); (3) studies must have group comparisons, such as medicated participants versus placebo/healthy controls/unmedicated participants; (4) studies must investigate the changes in the gut microbiome. There were no limitations regarding the existence of psychotic features, the presence of comorbid anxiety symptoms or disorders, or inadequate responses to prior treatments.

The exclusion criteria were as follows: (1) individual case reports relating to medications; (2) conference submissions that lack complete quantitative or qualitative reports; (3) studies that terminated early without efficacy analysis.

### Data collection and search strategy

Strategies to identify eligible studies included a systematic search of five electronic databases: Ovid MEDLINE, EMBASE (via Ovid), APA PsycINFO (via Ovid), PubMed and SCOPUS, up to 13 August 2024. The search was conducted with assistance from a library and information science specialist (LD) to ensure the inclusion of all relevant articles. The search included three concepts combined with boolean AND: bipolar disorder, gut microbiome, and mood stabilizing drugs. Each concept included subject headings and synonyms combined with Boolean OR and was optimized to run in each database. No methodological, publication, language or date limits were added to the search. The full search strategy is available in **S1 Table**.

All citations were reviewed by title, then by abstract, then by full text and were screened according to the inclusion and exclusion criteria by two reviewers independently (TAB and BRO) using the Covidence tool (194). When there was uncertainty regarding the inclusion of an article or disagreement, the final decision was made by AG. A total of 12 full-text articles were selected (**Fig 1**).

**Fig 1.**
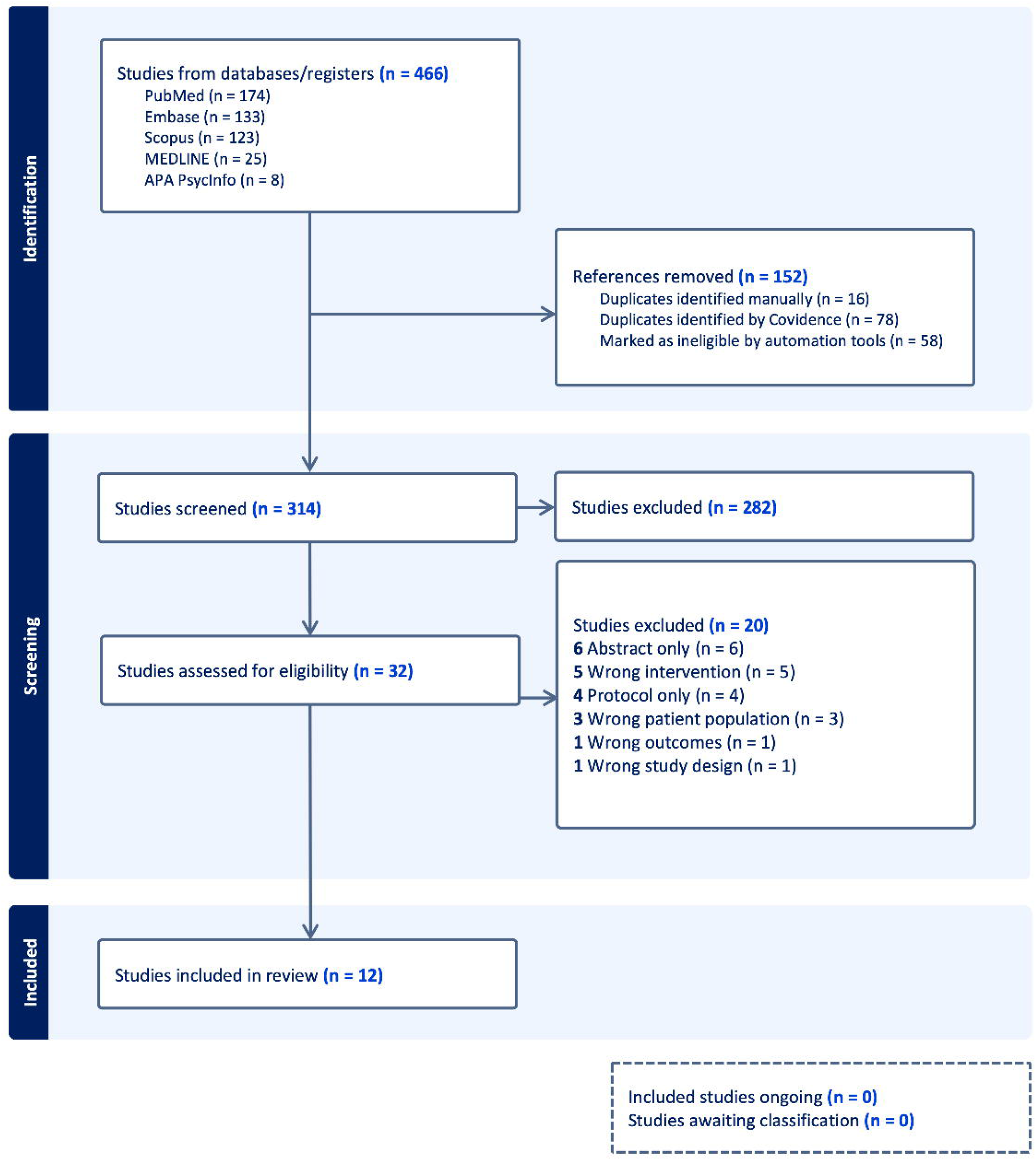
Study selection with PRISMA flow diagram.

### Data extraction

Two authors (TAB and BRO) systematically extracted the following information from the included articles into an Excel spreadsheet. Data were extracted in the following categories: (1) demographic and clinical characteristics of the study samples, including sample size, age range, sex ratios, BMI, diagnostic group, medications, inclusion criteria, and exclusion criteria; (2) microbiome analytic methodology includes gut genetic quantification, gut diversity measures, brain imaging (in selected studies), and covariates’ effects; (3) study findings, including group differences in taxonomic abundance and correlations between medications and associated gut microbiome characteristic changes. The primary outcomes from these studies were response to treatment and all-cause discontinuation (e.g., withdrawal consent). Since the methods and outcome measures in each study vary greatly from one another, we reported these differences and all details and did not have to do any data conversion.

### Risk of bias and certainty of evidence assessment

Given the diverse study designs in our systematic review, risk of bias in included studies was evaluated using the Cochrane risk of bias ROBINS-I V2 tool, which is suitable for cross-sectional interventional and longitudinal interventional studies (90). Two reviewers (TAB and BRO) independently assessed the studies for potential biases, including selection and reporting biases, and resolved discrepancies through discussion (**Fig 2**).

**Fig 2.**
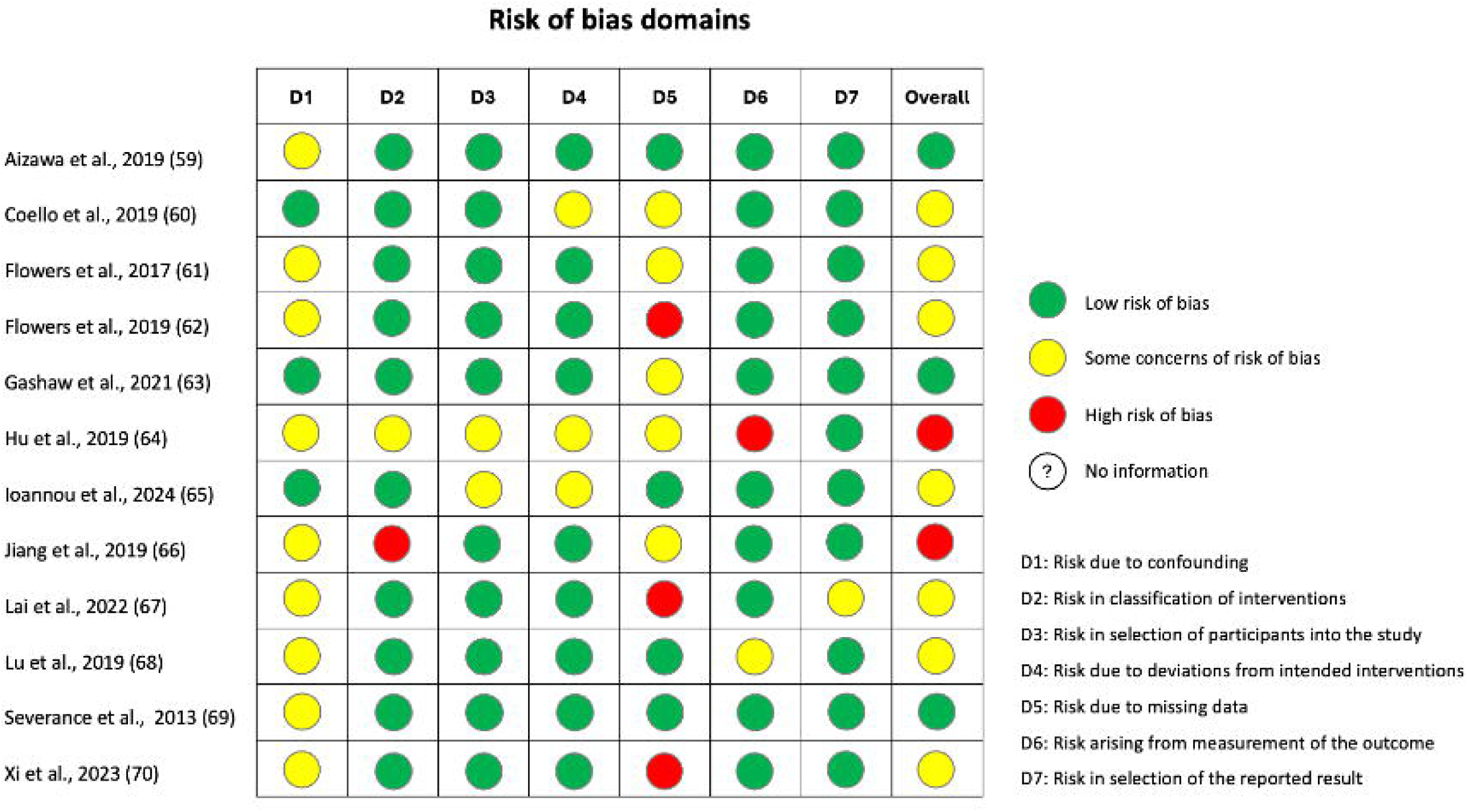
Risk of bias assessment.

The certainty of the evidence was assessed using the GRADE approach. This method evaluated study design, findings’ consistency, evidence’s directness, and effect estimates’ precision. The assessment provided an overall confidence rating for each outcome, guiding the interpretation of results and conclusions. A summary of findings table was created using the GRADE Working Group’s software GRADEpro GDT (**S2 Table**).

## Results

The search strategy is provided in **S1 Table**. **Fig 1** illustrates the literature search process. Database searches identified 466 references; 314 articles were identified from all databases after removing duplicates. Of these, 282 were excluded based on the title and abstract and 20 based on the full text, resulting in 12 articles for inclusion (**Table 1**).

**Table 1.** Studies included for systematic review.

| Author | Name |
| --- | --- |
| Aizawa et al., 2019 (91) | Bifidobacterium and lactobacillus counts in the gut microbiota of patients with bipolar disorder and healthy controls |
| Coello et al., 2019 (92) | Gut microbiota composition in patients with newly diagnosed bipolar disorder and their unaffected first-degree relatives |
| Flowers et al., 2017 (93) | Interaction between atypical antipsychotics and the gut microbiome in a bipolar disease cohort |
| Flowers et al., 2019 (94) | Effects of atypical antipsychotic treatment and resistant starch supplementation on gut microbiome composition in a cohort of patients with bipolar disorder or schizophrenia. |
| Gashaw et al., 2021 (95) | Assessment of gut bacteria profile and antibiotic resistance pattern among psychotropic drug users: comparative cross-sectional study |
| Hu et al., 2019 (96) | Gut microbiota changes in patients with bipolar depression |
| Ioannou et al., 2024 (97) | Reproducible gut microbial signatures in bipolar and schizophrenia spectrum disorders: a metagenome-wide study |
| Jiang et al., 2019 (98) | The microbiome in bipolar depression: a longitudinal study of one pair of monozygotic twins |
| Lai et al., 2022 (99) | Metagenomic analysis reveals gut bacterial signatures for diagnosis and treatment outcome prediction in bipolar depression |
| Lu et al., 2019 (100) | Gut microbiota in bipolar depression and its relationship to brain function: an advanced exploration. |
| Severance et al., 2013 (101) | Discordant patterns of bacterial translocation markers and implications for innate immune imbalances in schizophrenia. |
| Xi et al., 2023 (102) | Brain-gut microbiota multimodal predictive model in patients with bipolar depression |

### Overview of study characteristics

Demographic and clinical characteristics of the study samples are presented in detail in **Table 2** and **3**. The study with the largest sample was conducted in Baltimore, U.S. (N = 294), and the remaining study samples ranged from N = 6 to N = 231. All studies were conducted between 2013 and 2024. Location varied widely between studies, including the U.S., China, Denmark, Ethiopia, Japan, and the Netherlands, and most patients were recruited from local psychiatric hospitals. In some studies, a variety of medication combinations was used, such as antidepressants and benzodiazepines, further complicating the results. Most studies (10/12) include HCs (**Table 2**). In two studies, BP patients were further divided into quetiapine-responded and non-responded patients based on whether the treatment reduced HAMD-17 score by 50% compared to baseline and a random forest classification model (99,102). Two studies also recruited unaffected relatives (UR) and BP patients (92,98). Three studies also included patients with schizophrenia spectrum disorder (SSD) (95,97,101).

**Table 2.** Demographic and clinical characteristics of included studies.

| Study | Sample size* | Location | Design | Sample type | Diagnosis | Response | Medications (N) |
| --- | --- | --- | --- | --- | --- | --- | --- |
| Aizawa 2019 (91) | N = 122<br>HC = 58 (♀ 36; ♂ 22)<br>BP = 39 (♀ 22; ♂ 17)<br>AAP = 13<br>LL = 29 | Tokyo, Japan | Cross-sectional observational cohort | Inpatient<br>Community | BP = 39<br>BP-I = 13<br>BP-II = 26 | N/A | AAP: Sodium valproate (8), Carbamazepine (4)<br>Antidepressant (12)<br>Lithium (16)<br>Lamotrigine (13) |
| Coello 2019 (92) | N = 229<br>HC = 77 (♀ 47; ♂ 30)<br>UR = 39 (♀ 21; ♂ 18)<br>BP = 113 (♀ 70; ♂ 43) | Copenhagen, Denmark | Cross-sectional observational cohort | Community | BP-I = 44<br>BP-II = 65<br>BP NOS = 4 | N/A | AAP (43)<br>Antidepressants (30)<br>Antiepileptics (58)<br>Lithium (44) |
|  | AAP = 43<br>LL = 44 |  |  |  |  |  |  |
| Flowers 2017<br>(93) | N = 117**<br>AAP = 49 (♀ 34; ♂ 12)<br>NAAP = 68 (♀ 48; ♂ 21)<br>LL = 31 | Michigan, U.S. | Cross-sectional<br>observational<br>cohort | Community | BP = 117 | N/A | **AAP:** Clozapine, Olanzapine, Risperidone, Quetiapine, Asenipine, Ziprasodone, Lurasidone, Aripiprazole, Paliperidone, Iloperidone.<br>**Antidepressants** (52): Bupropion, Venlafaxine, Sertraline, Duloxetine, Fluoxetine, Citalopram, Escitalopram.<br>**Mood stabilizers** (60): Topiramate, Phenobarbital, Lamotrigine, Gabapentin, Divalproex sodium, Carbamazepine, Lithium.<br>**Benzodiazepines** (32): Lorazepam, Alprazolam, Temazepam, Clonazepam, Diazepam. |
| Flowers 2019<br>(94) | N = 37<br>AAP = 21 (♀ 9; ♂ 12)<br>LL = 16 (♀ 7; ♂ 9) | Michigan, U.S. | Cross-sectional<br>interventional<br>longitudinal<br>cohort | Community | **AAP group:**<br>BP-I = 9<br>BP-II = 3<br>SZA = 5<br>SCZ = 4<br>**LL group:**<br>BP-I = 10<br>BP-II = 6 | N/A | **AAP (3 weeks):** Clozapine, Olanzapine, Risperidone, Quetiapine, Ziprasidone + Starch (2 weeks).<br>**Mood stabilizer (3 weeks):** Lithium, Lamotrigine<br>**Concomitant medications:** Antidepressants, Benzodiazepines, Medications for hypertension, diabetes mellitus, and hyperlipidemia. |
| Gashaw 2021<br>(95) | N = 214**<br>HC = 107 (♀ 27; ♂ 80)<br>SCZ = 57<br>BP = 28<br>Depression = 21 | Jimma,<br>Ethiopia | Cross-sectional<br>observational<br>cohort | Inpatient<br>Community | SCZ = 57<br>BP = 28<br>Depression = 21 | N/A | **Antipsychotics** (54)<br>**Mood stabilizers** (14)<br>**Antidepressants** (11)<br>**Combination therapy** (28) |
| Hu 2019<br>(96) | N = 97<br>HC = 45 (♀ 22; ♂ 23)<br>BP = 52 (♀ 25; ♂ 27) | Zhejiang,<br>China | Cross-sectional<br>interventional<br>longitudinal<br>cohort | Inpatient<br>Community | BP-I = 12<br>BP-II = 38<br>BP NOS = 2 | N/A | **Quetiapine** (4 weeks)<br>Maintenance dose: 200-300mg/day |
| Ioannou 2024<br>(97) | N = 231<br>HC = 128 (♀ 77; ♂ 51)<br>BP = 66 (♀ 48; ♂ 18)<br>AAP = 34<br>LL = 35<br>NAAP = 35<br>SSD = 37 (♀ 16; ♂ 21)<br>AAP = 28<br>LL = 3 | Groningen,<br>Netherlands | Cross-sectional<br>observational<br>cohort | Inpatient<br>Community | BP = 66<br>SCZ/SZA = 37 | N/A | **Antipsychotics** (62)<br>**Lithium** (38)<br>**Antidepressants** (29)<br>**Anticonvulsants** (23) |
|  | NAAP = 15 |  |  |  |  |  |  |
| Jiang 2019<br>(98) | N = 6<br>HC = 4 (♀ 2; ♂ 2)<br>UR = 1 (♀ 1)<br>BP = 1 (♀ 1) | Zhejiang,<br>China | Longitudinal<br>case-control | Inpatient<br>Community | BP = 1 | N/A | **Lamotrigine**<br>Maintenance dose: 100mg/day<br>**Quetiapine**<br>Maintenance dose: 50mg/day |
| Lai 2022<br>(99) | N = 122<br>HC = 60 (♀ 31; ♂ 29)<br>BP = 62 (♀ 23; ♂ 39) | Zhejiang,<br>China | Cross-sectional<br>interventional<br>longitudinal<br>cohort | Inpatient<br>Community | BP-I = 12<br>BP-II = 45<br>BP NOS = 5 | NBP = 19***<br>RBP = 35<br>Other = 9 | **Quetiapine** (4 weeks)<br>Initial dose: 50mg/day<br>Maintenance dose: 200-300mg/day |
| Lu 2019<br>(100) | N = 63<br>HC = 27 (♀ 12; ♂ 15)<br>BP = 36 (♀ 15; ♂ 21) | Zhejiang,<br>China | Cross-sectional<br>interventional<br>longitudinal<br>cohort<br>RCT | Inpatient<br>Community | BP-I = 10<br>BP-II = 26 | N/A | **Quetiapine** (4 weeks)<br>Initial dose: 50mg/day<br>Maintenance dose: 300mg/day<br>**Propranolol**: 10-20mg<br>**Glycerine enema** |
| Severance<br>2013 (101) | N = 294<br>HC = 78 (♀ 56; ♂ 22)<br>BP = 75 (♀ 52; ♂ 23)<br>AAP = 57<br>NAAP = 18<br>SCZ = 141 (♀ 56; ♂ 85) | Baltimore,<br>U.S.<br>Cologne,<br>Germany | Cross-sectional<br>observational<br>cohort | N/A | BP = 75<br>SCZ = 141 | N/A | **Antipsychotics**<br>SCZ = 141<br>BP = 57 |
| Xi 2023<br>(102) | N = 142<br>HC = 39 (♀ 23; ♂ 16)<br>BP = 103<br>QTP = 75 (♀ 38; ♂ 37) | Zhejiang,<br>China | Cross-sectional<br>interventional<br>longitudinal<br>cohort | Inpatient<br>Community | BP = 103 | RBP = 43***<br>NBP = 32 | **Quetiapine** (4 weeks)<br>Maintenance dose: 200-300mg/day |

AAP, atypical antipsychotics.
BP, bipolar disorder.
HC, healthy controls.
LL, lithium- or lamotrigine-treated group.
NAPP, No atypical antipsychotics.
NBP, quetiapine non-responded patients.
NOS, not otherwise specified.
QTP, quetiapine.
RBP, quetiapine responded patients.
RCT, registered clinical trial.
SCZ, schizophrenia.
SSD, schizophrenia spectrum disorders.
SZA, schizoaffective disorder.
UR, unaffected first-degree relative.

### Study design

Most studies employed a cross-sectional cohort design. There was significant heterogeneity between studies; among the 12 articles, one was a registered clinical trial (100) and six were longitudinal studies (94,96,98–100,102); thus only a subset shared comparability. Three studies specified a priori taxa for study, as opposed to studying the entire gut microbiota (95,100,101). Four studies (4/12) were interventional, where participants underwent a 3-4 week treatment with psychotropic medications (94,99,100,102). These studies employed a fixed dose of quetiapine (300mg/day) without a discernible dose-response correlation. The remaining eight studies were observational, where researchers monitored and compared the effects of psychotropic medications between individuals who were taking psychotropics and those who did not.

Potential confounds when studying the microbiome, including age, sex, BMI, and medications, were all recorded. BP patients were recruited and diagnosed using various means, including admission in the hospital’s inpatient/outpatient unit, using DSM-IV or DSM-V, Young Mania Rating Scale (YMRS), Hamilton Anxiety Rating Scale (HAMA), Hamilton Depression Rating Scale (HDRS), Montgomery-Asberg Depression Rating Scale (MADRS), Mini International Neuropsychiatric Interview (MINI), Diagnostic Interview for Genetic Studies (DIGS). Most studies excluded patients with substance abuse, cognitive impairments, intellectual disabilities, pregnancy, severe physical disabilities, and uses of antibiotics, probiotics, and/or prebiotics within 4 weeks of screening. A variety of sequencing methods (metagenomic sequencing, 16s RNA sequencing, qPCR) and analysis pipelines (PERMOVA, AMOVA, LEfSe, PCoA) were used to characterize microbiome composition (**Table 3**).

**Table 3.**
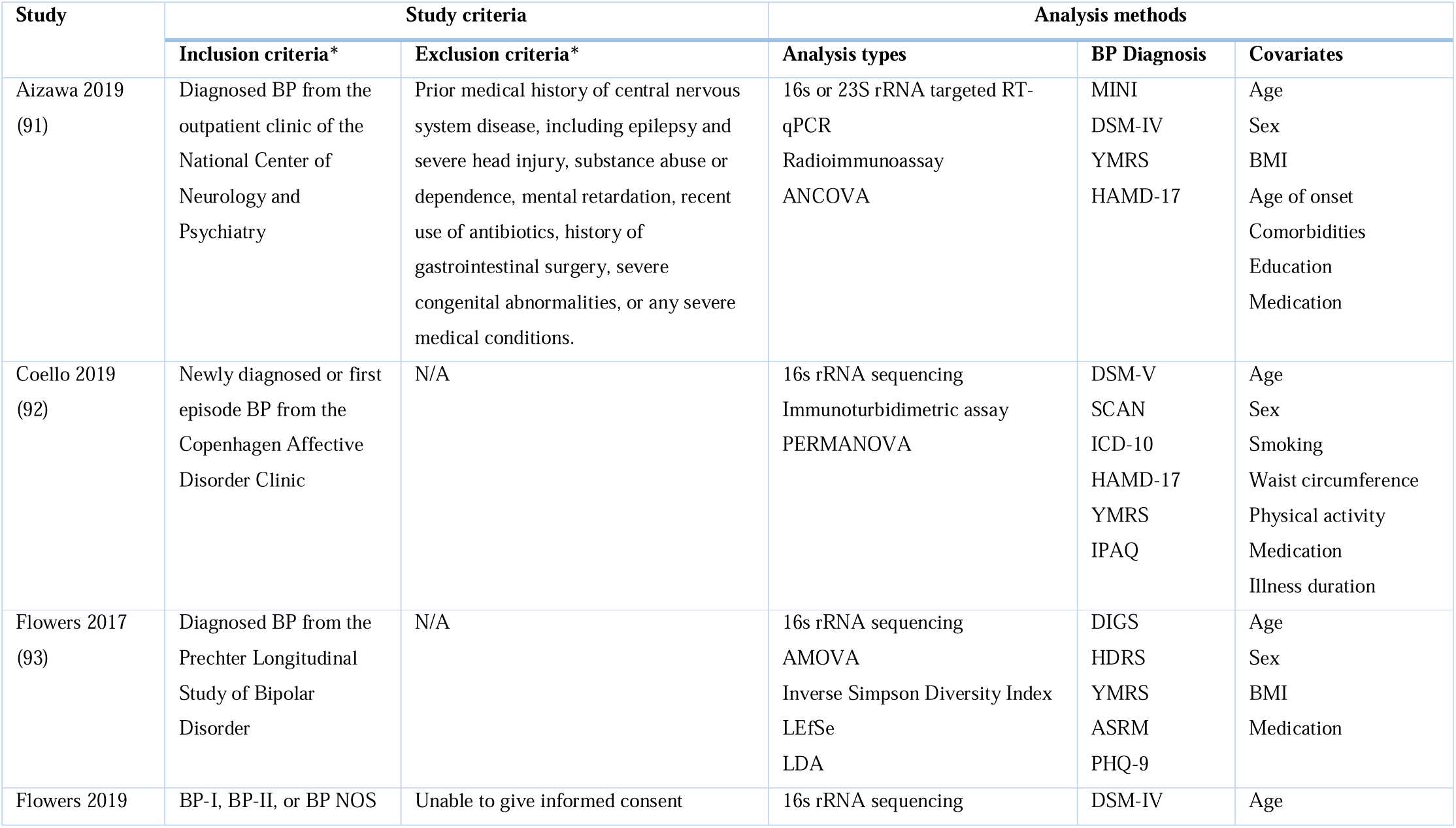
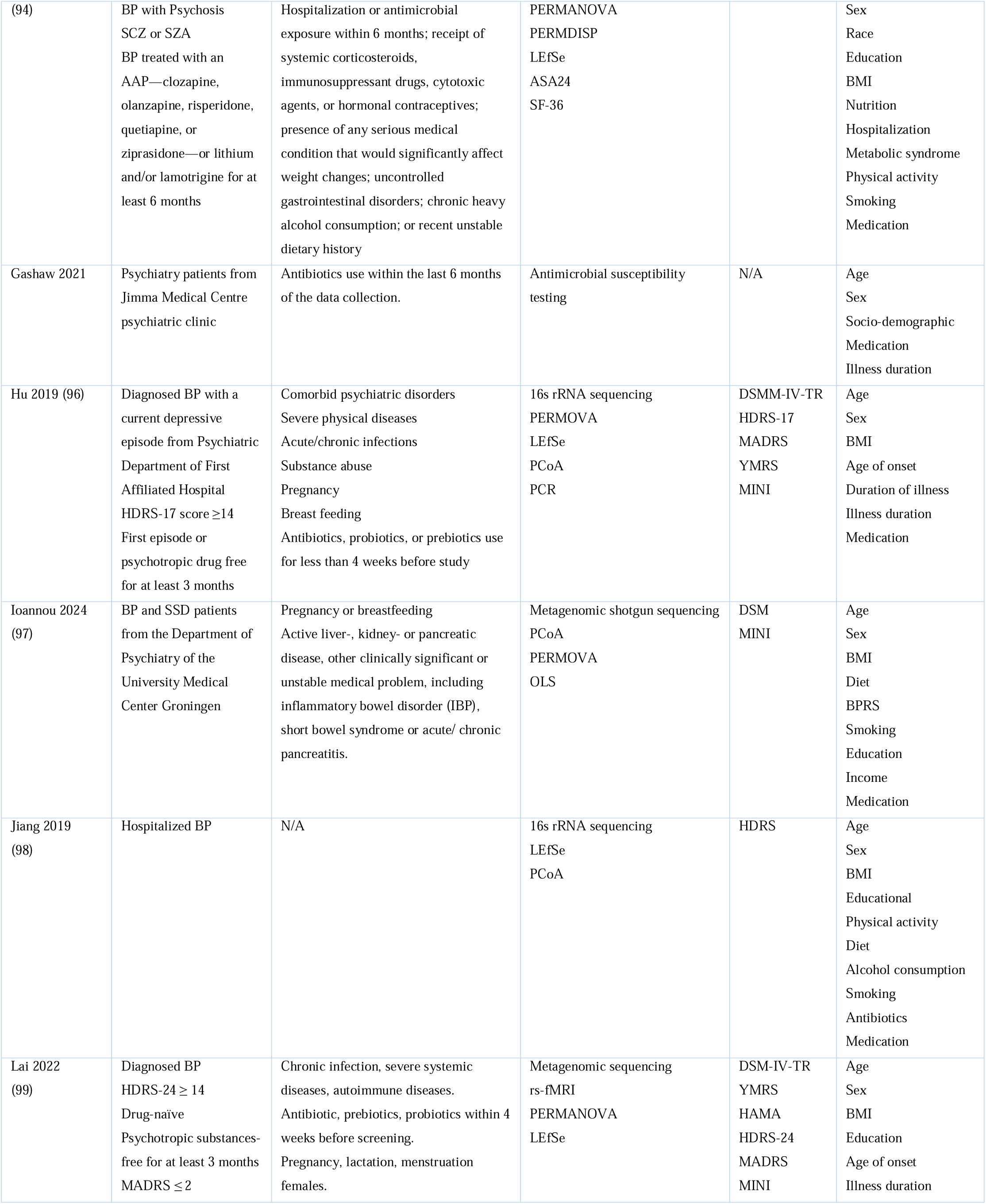
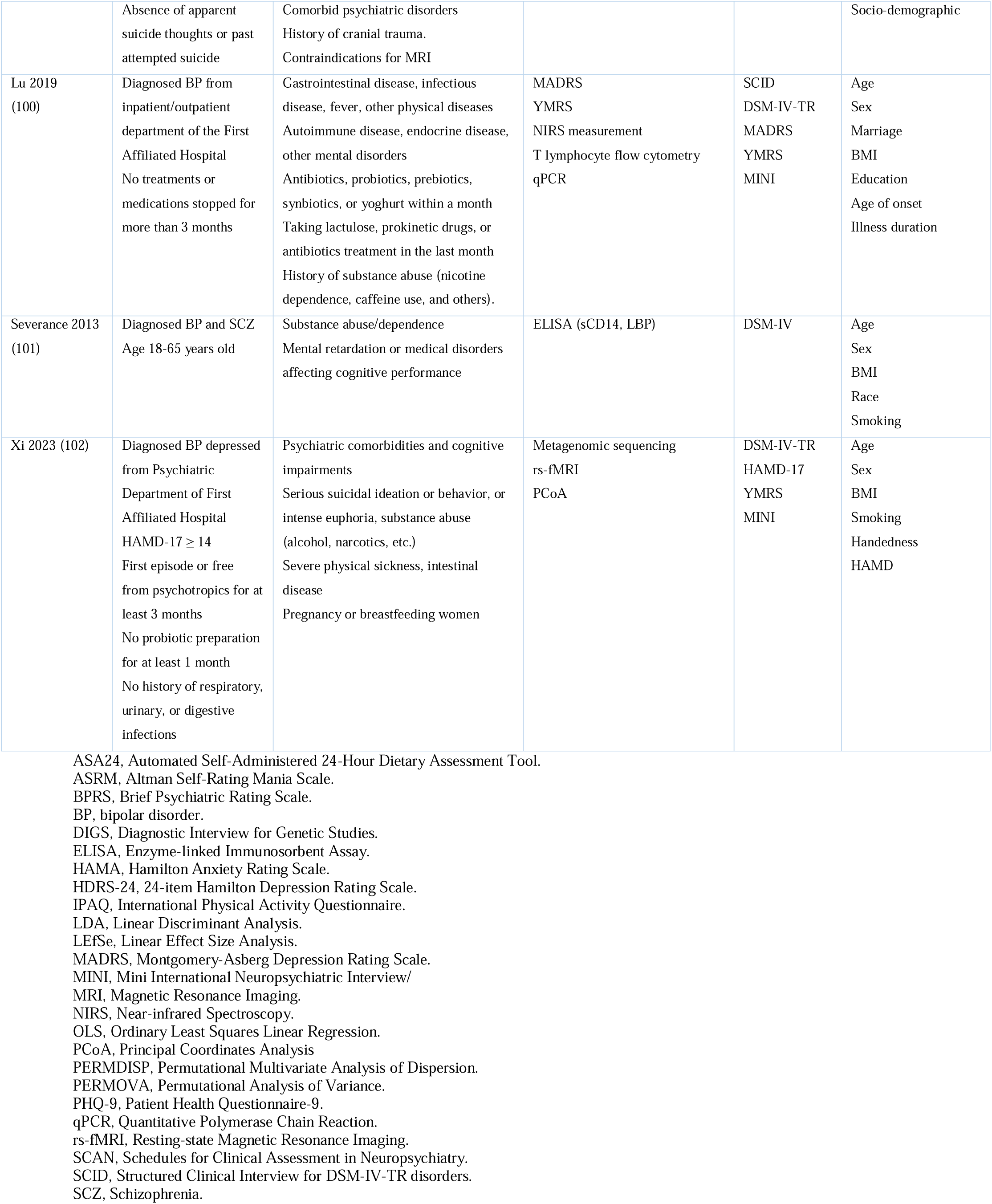
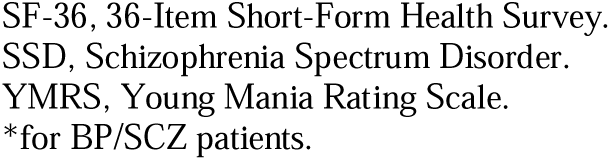
Study criteria and analysis methodology of included studies.

### Quality assessment results

The quality assessment results are reported in **Fig 2** and **S2 Table**. ROBINS-I V2 tool was used to evaluate the risk of bias for all included studies, which examines seven bias domains: confounding, intervention classification, participant selection, deviations from intended interventions, missing data, outcome measurement, and result reporting. There was author overlap between papers, with the same groups having published most papers (i.e., Flowers et al. (93,94) and Hu et al. (96,99,100,102)), which might create overlap between study populations. We did not exclude these studies as we considered that the risk of bias would be negligible. Most studies acknowledged their limitations due to the lack of control for confounding factors, such as diet and lifestyle (**Fig 2**). We also assessed the certainty of evidence using the GRADE approach for any reported correlation between psychotropic use and changes in gut microbial species (**S2 Table**) (103). Evidence is categorized by high, moderate, low and very low certainty.

### Effects of mood stabilizers on gut microbiome in bipolar disorder

Our collected research studies yield mixed results. In two studies, based on the degree of improvement in depression severity post-treatment (quetiapine) compared to baseline (HAMD-17/HDRS-124 change ≥ 50%), patients were classified into responder and non-responder groups, with the responders’ scores significantly lower than non-responders (99,102). Interestingly, before quetiapine treatment, non-responders had significantly lower gut microbial alpha diversity than responders, with *Eubacterium biforme*, *Weissella confusa, Oribacterium sinus*, and *Barnesiella intestinihominis* being more abundant in responders and *Clostridium bartlettii*, *Bacteroides_sp_2_1_22*, and *Bacteroides_sp_3_1_19* more in non-responders (102).

There were generally no significant associations between alpha or beta diversity and psychotropic treatment (91,94,97,100,102). However, in two studies, Lai et al., 2022 (99) found increased alpha diversity in BP patients after a 4-week treatment of atypical antipsychotics (AAPs), and the Flowers research group noticed a significantly decreased alpha diversity in treated female patients that was not observed in males (93,94). In a longitudinal monozygotic twin study, microbiome abundance was also at the lowest level following a female patient’s hospitalization and treatments of lamotrigine, lithium, and quetiapine (98). This sex-dependent change is consistent with preclinical studies of AAPs-treated rat models like olanzapine and risperidone, where weight gain and microbial composition changes only occurred in females (104–106). Olanzapine was shown to convert gut microbial composition into an obesogenic profile (78) and risperidone seemed to decrease “lean gut microbiota” (107). Children and adolescents under chronic risperidone treatment also showed an upregulation of metabolic pathways linked with weight gain (108). These findings reinforce that AAPs alter the inflammatory milieu and metabolic profile of BP patients drastically, shifting them towards an obesogenic one.

The emergence of antimicrobial resistance among gut bacteria in patients treated with psychotropic medications was a notable concern. The most frequently isolated bacteria from BP patients and HC were *E. coli*, with antibiotic resistance patterns differing between the two groups (95). Amoxicillin clavulanic acid, cefuroxime, ceftriaxone, cefepime, cefotaxime, meropenem, ciprofloxacin, and tetracycline resistance were significantly higher among *E. coli* bacteria isolated from participants taking psychotropic drugs than *E. coli* bacteria isolated from apparently HC. This shows that multidrug-resistant bacteria and extended-spectrum beta-lactamase-producing (ESBL) strains are significantly more prevalent in BP patients taking psychotropics. The overall rate of multidrug resistance among bacteria isolated from patients taking psychotropics was 63%, whereas it accounts for 42.1% among bacteria isolated from HC. Furthermore, the odds of isolating ESBL-producing Enterobacteriaceae were notably higher in patients treated with antipsychotic drugs for over a year. This raises questions about the indirect effects of psychotropic medications on microbial ecology, possibly mediated by altered gut environments or medication-induced immune modulation (95).

While some studies reported changes only at the genus level, others also documented changes at the species level. When analyzing the changes by specific bacteria, we identified three groups, encompassing both genus and species: bacteria that increased in abundance after treatment, those that did not, and those with mixed results (**Table 4**).

**Table 4.**
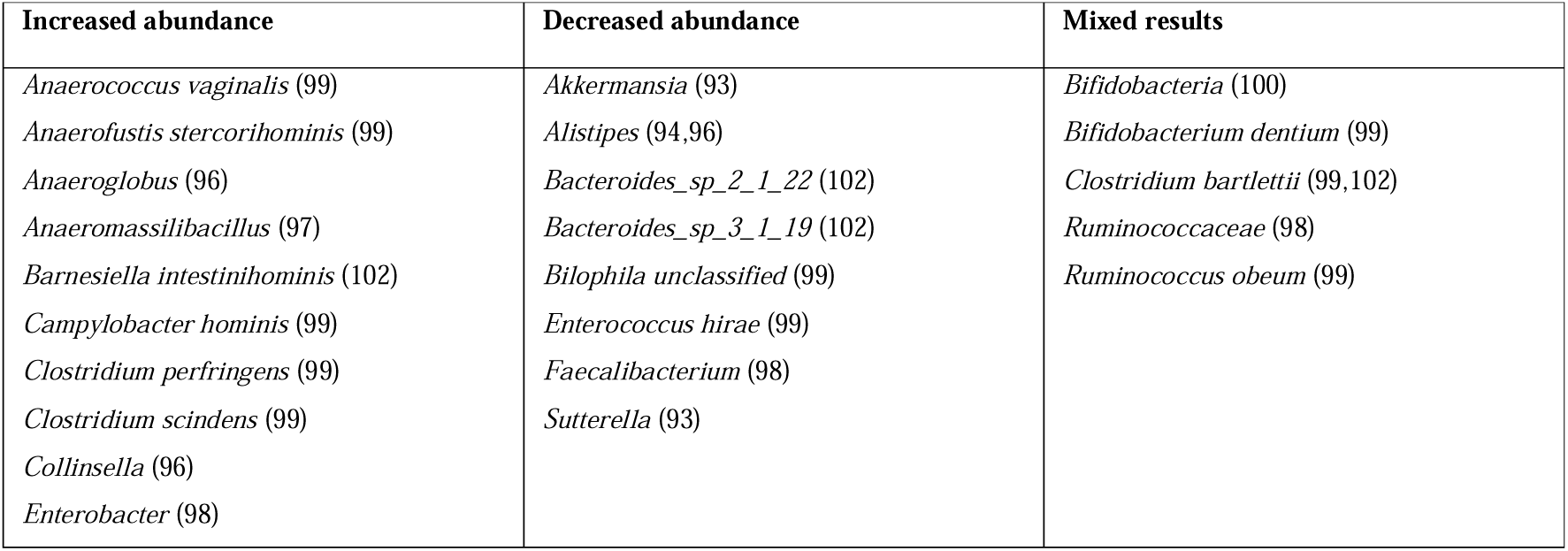
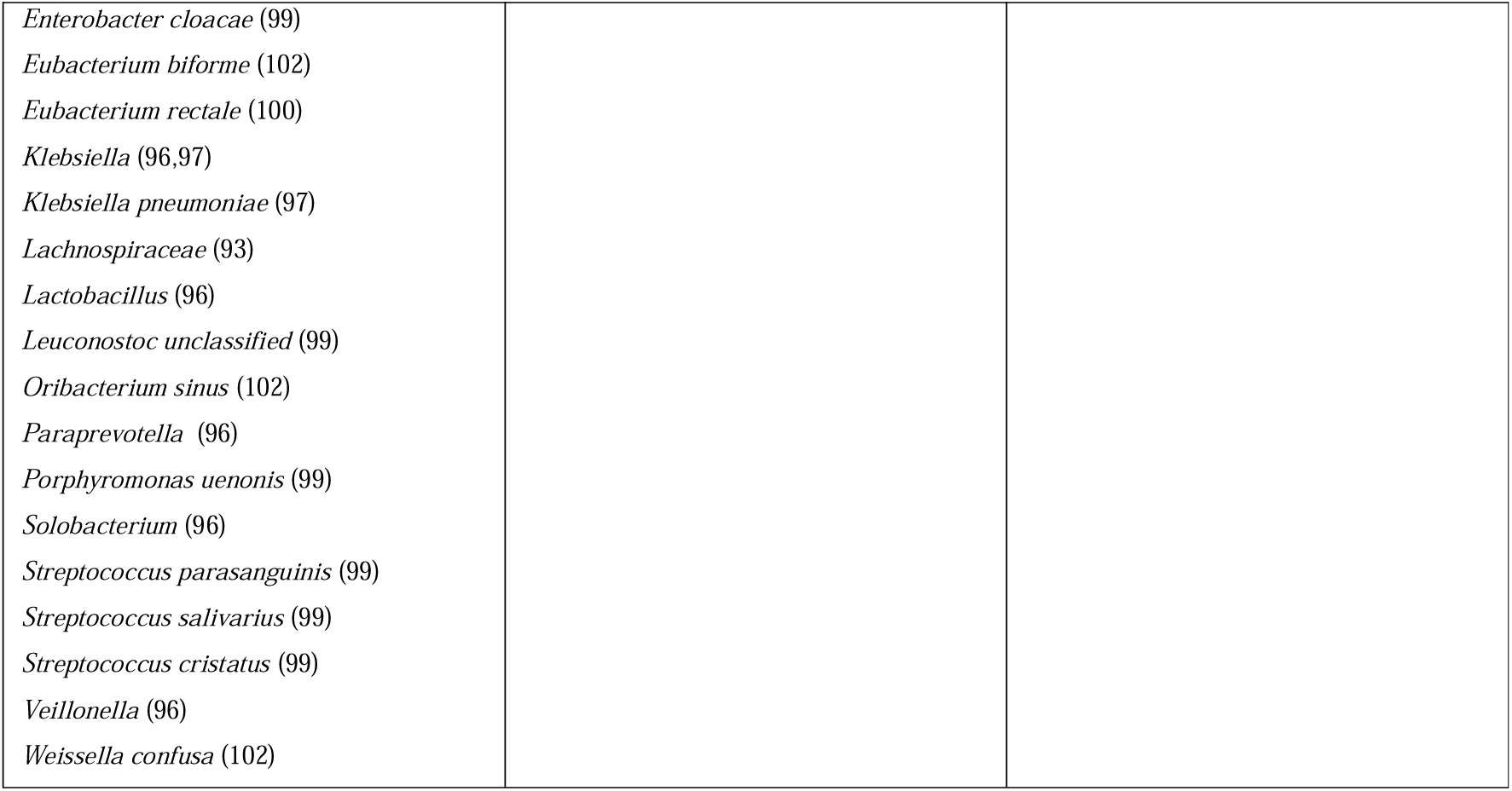
Changes in gut microbiome composition upon psychotropics treatment.

### Increased bacteria abundance post-treatment

Increased abundance of 23 genera and 18 bacterial species were observed in BP patients treated with antipsychotics and lithium, and anticonvulsant and antidepressant use were not associated with any of the taxa (**Table 4**). These specific species include *Anaerococcus vaginalis, Anaerofustis stercorihominis, Barnesiella intestinihominis, Campylobacter hominis, Clostridium perfringens, Clostridium scindens, Enterobacter cloacae, Eubacterium biforme, Eubacterium rectale, Klebsiella pneumoniae, Leuconostoc unclassified, Oribacterium sinus, Porphyromonas uenonis, Ruminococcus obeum, Streptococcus parasanguinis, Streptococcus salivarius, Streptococcus cristatus, Weissella confusa.* These encompass a diverse range of species, including both Gram-positive and Gram-negative bacteria. Many Gram-positive species, such as *Lactobacillus*, are well-known for their roles in human microbiota, especially in the gut, and are involved in processes like fermentation, digestion, and the maintenance of gut health (109,110). *Eubacterium biforme*, *Eubacterium rectale*, and *Ruminococcus obeum* produce SCFAs like butyrate, maintaining gut barrier integrity and reducing inflammation . Their increased abundance could indicate adaptive microbial changes or therapeutic effects aimed at restoring gut homeostasis in treated patients.

*Clostridium Cluster IV* is a butyrate-producing bacterium (117) and belongs to beneficial bacteria (100). However, some of these bacteria are pathogenic and can cause severe infection. *Clostridium perfringens* is an enteric pathogen that can produce various toxins and hydrolytic enzymes associated with intestinal diseases in humans and animals (99,118). Gram-negative bacteria like *Campylobacter hominis* and *Klebsiella pneumoniae* are also notable for their clinical significance; *Klebsiella pneumoniae* are often opportunistic pathogens implicated in the urinary tract (119,120), while *Campylobacter hominis* is a known pathogen in gastrointestinal diseases (121,122).

In a longitudinal monozygotic twin study, KEGG pathway level 3 lipopolysaccharides (LPS) biosynthesis genes were over-represented in the gut following hospitalization and treatments of lamotrigine, lithium, and quetiapine (active state) (98). LPS are bacterial surface glycolipids produced by Gram-negative that can stimulate inflammation in the gut microbiome (123). During the remissive state, LPS biosynthesis genes are underrepresented compared to the active stage of BP, leading the patient’s gut microbiome to become more similar to that of their monozygotic healthy twin. Throughout the responsive state (discharge and lithium withdrawal) and remissive (outpatient visit) BP state, *Ruminococcaceae* and *Faecalibacterium* abundance also increased significantly while *Enterobacter* declined (98). This is consistent with previous studies on the general effects of lithium on the gut microbiome (79,124). These results suggest that gut microbial function partially recovered after patients achieved full remission. Moreover, the overrepresented LPS biosynthesis genes may be related to the overgrowth of *Enterobacter* during the active stage, and patients’ gut microbiome compositions corrected as they moved towards a healthy status (98).

### Decreased bacteria abundance post-treatment

Following a 4-week quetiapine treatment, the abundance of *Akkermansia* (93), *Alistipes* (94,96), *Bacteroides_sp_2_1_22* (102), *Bacteroides_sp_3_1_19* (102), *Bilophila unclassified* (99), *Enterococcus hirae* (99), *Faecalibacterium* (98), *Ruminococcaceae* (98), *Sutterella* (93)*. Alistipes* bacteria are within the *Bacteroidetes* phylum, enriched in untreated BP patients (96), and inversely associated with obesity and an animal-rich diet (125,126). *Akkermansia* was significantly decreased in non-obese patients treated with AAPs (93). This genus was previously inversely correlated with inflammation, insulin resistance, and lipid metabolism (127). Nevertheless, in two studies, no significant difference was found in bacterial counts before and after treatment (91,92).

### Mixed findings

The abundance of *Clostridium bartlettii* varied across two studies (99,102) (**Table 4**). Both studies classified the BP patients as quetiapine responders and non-responders after a four-week treatment based on HAMD changes. Lai et al., 2022 identified an increased abundance of *Clostridium bartlettii* following quetiapine treatment in the study. This was accompanied by improvement in depressive symptoms following treatment, which appears to correlate with crucial emotion and behaviour recovery in BP patients but increased metabolic dysfunction (99).

Conversely, Xi et al. 2023, from the same research group, observed elevated *Clostridium bartlettii* levels in quetiapine non-responders, linking it to 5-HT deficiency and poorer remission outcomes (102). They argued that as that species has been linked to 5-HT deficiency (128), patients with excess *Clostridium bartlettii* may not do well in remission post-quetiapine treatment (102). They also added that *Clostridium bartlettii* had been associated with increased weight gain (99,129), with a decreased fecal abundance of *Clostridium bartlettii* following metformin treatment in three trials (130–132). These findings suggest that the species may have a complex role in quetiapine treatment responses, potentially influencing metabolic dysfunction, weight gain, and mood regulation. However, whether and how the species has detrimental or rescuing effects on BP symptoms is unclear. Psychotropics (particularly quetiapine) may partially improve mood symptoms through interactions with beneficial bacteria while increasing the risk of metabolic disturbance in more susceptible patients through interactions with other species of gut bacteria.

The findings for *Bifidobacteria* were also inconsistent. One study reported a significant increase in *Bifidobacteria* counts (100), whereas another found a decrease in *Bifidobacterium dentium* after quetiapine treatment (99) (**Table 4**). In a study conducted by Lu et al., after a four-week quetiapine treatment, the populations of both anaerobic *Bifidobacteria* and *Eubacterium rectale* proliferated compared to baseline, coinciding with a decline in the MADRS score and an increase in *Bifidobacteria-to-Enterobacteriaceae* (B/E) ratio (100). The authors suggested these bacteria could serve as biomarkers for therapeutic effects, but their lack of a placebo control leaves causality unclear (**Fig. 2**). Interestingly, in a separate study conducted by the same group, a four-week quetiapine treatment decreased the abundance *Bifidobacterium dentium*, implying its potential role in improved depressive symptoms post-treatment (99). Nevertheless, the absence of healthy controls in their fMRI analysis presents some biases and limits these associations (**Fig 2**).

The findings on the abundance of *Ruminococcus obeum* species and the family *Ruminococcaceae* following four weeks of quetiapine treatment present another conflict. While one study shows an increase in *Ruminococcus obeum* following quetiapine treatment (97), another identifies an ongoing increase in *Ruminococcaceae* during the responsive and remissive states of BD, despite initial decreases in the active state (98). This is particularly intriguing given that this same genus was enriched in the HC group. This warrant further investigation into the mechanistic pathways of the bacteria and their metabolites in BD progression and therapy.

### Correlated functional imaging changes

Two studies from our search employed fMRI to investigate the correlation between gut bacterial species and neural activity (99,102). However, Lai et al., 2022 only collected fMRI results from untreated BP patients (99). In contrast, Xi et al., 2023 collected fMRI data from HC and all BP patients before and after they underwent a 4-week quetiapine treatment (102). They showed that the FC and grey matter volume (GMV) of quetiapine-responding BP patients were closer to those of HC than non-responders, suggesting that successful quetiapine treatment may restore some neural function and connectivity to a more normative state. Moreover, quetiapine-responding patients had lower intra-network FC within DMN and between the DMN and other networks (i.e., frontoparietal network and ventral attention). This lower FC was negatively correlated with *TM7_TM7b* (bacteria from the *Saccharibacteria* phylum)*, Weissella confusa,* and positively correlated with *Clostridium bartlettii*. FC within the DMN is positively correlated with 5-HT signal transduction (133), which is related to the primary action of quetiapine (e.g., blocking dopamine D2 (134) and 5-HT_2A_ receptors (135,136), while activating 5-HT_1A_ (137)). Significantly higher DMN connections indicate that non-responders have dysfunctional or abnormal 5-HT signal transduction, making 5-HT more difficult for the quetiapine to bind to the 5-HT receptor (102) (**Fig 3**).

**Fig 3.**
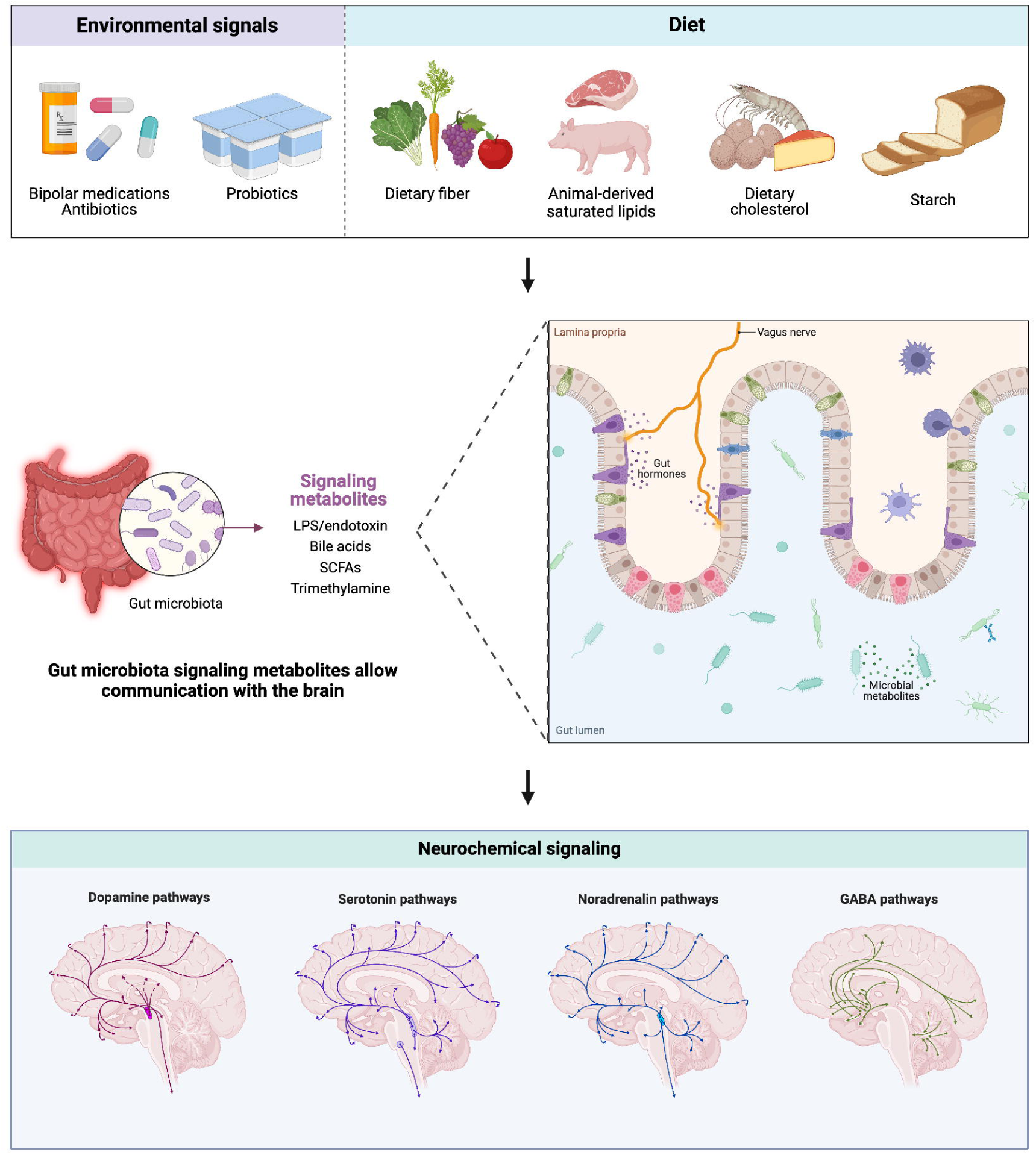
Proposed effects of environmental factors on the gut microbiome composition and neurochemical signalling in bipolar disorder.

These imaging findings suggest that quetiapine non-responders may have additionally abnormal 5-HT signaling, leading to insufficient quetiapine binding at key receptors. This dysfunction within the 5-HT system contributes to the persistent DMN hyperconnectivity observed, amplifying an already maladaptive neural state.

## Discussion

The gut-brain axis is a bidirectional communication system involving the CNS, enteric nervous system, and gut microbiota. The gut microbiome interacts with the brain via neurotransmitters, metabolites, and hormones (138). Thus, changes to the CNS in psychiatric disorders, such as neurotransmitter dysregulations, can affect the gastrointestinal microenvironment and vice versa (139). Studying the effects of medications on the gut microbiome in BP is essential for advancing our understanding of treatment mechanisms and improving patient outcomes. A significant number of individuals with BP do not respond adequately to existing medications such as mood stabilizers, antipsychotics, or antidepressants (140,141). Gut dysbiosis may perpetuate systemic inflammation or disrupt neurochemical signalling, such as serotonin and GABA pathways, contributing to poor treatment outcomes. By examining how medications influence gut bacteria, researchers can uncover underlying mechanisms of treatment resistance and identify new personalized strategies to support patients who do not respond to conventional therapies. The gut microbiome is highly individualized and shaped by diet, lifestyle, genetics, and environment (142). Medications may impact each person’s microbiome differently, contributing to variability in therapeutic outcomes. By studying these effects, researchers can develop microbiome-based biomarkers to predict how a patient will respond to specific treatments, enabling clinicians to tailor therapies to individual needs. This approach aligns with the growing emphasis on precision medicine in mental health care (143).

This review provides the most comprehensive analysis to date on the effects of BP treatments on the gut microbiome. We found that quetiapine and lithium were associated with an increased abundance of various gut bacteria species. Specifically, elevated levels of *Clostridium perfringens* and *Enterobacter cloacae*—opportunistic pathogens—suggest gut dysbiosis following treatment. On the other hand, the increased abundance *of Eubacterium biforme, Eubacterium rectale*, and *Ruminococcus obeum,* which produce beneficial SCFAs, indicates that quetiapine may have some positive effects on restoring gut homeostasis in treated patients. Additionally, the use of psychotropic drugs may contribute to antibiotic resistance, as observed in studies of *E. coli* from patients on these medications. This suggests that psychotropic drugs could promote the emergence of multidrug-resistant bacteria. Some studies also propose that gut microbiome composition could serve as a non-invasive biomarker for diagnosing and predicting treatment outcomes in mental health disorders like BP.

This review presents several limitations. We restricted the scope of this review to studies published in peer-reviewed journals, which introduces a potential for publication bias. Studies reporting significant or positive findings are more likely to be published, while those presenting null or negative results may remain unpublished. This bias could distort the conclusions regarding the effects of medications on the gut microbiome. Incorporating grey literature, such as conference proceedings or preprints, could mitigate this limitation by providing a more comprehensive and balanced representation of the available evidence. Such sources often include novel or preliminary findings, which are particularly valuable in rapidly evolving fields like microbiome research. Moreover, this review did not conduct any statistical analyses due to the substantial heterogeneity in the methodologies of the included studies. Variations in study design, populations, analytical techniques, and reporting standards precluded meaningful quantitative synthesis, further limiting the robustness of the conclusions.

The majority of included studies focus on the effects of specific medications like quetiapine and lithium, potentially neglecting other treatments used in BP, such as valproate. However, we include studies with negative findings (91). This limited scope might hinder the generalizability of findings to all BP medications. Studies assessing the gut microbiome and BP treatments likely vary in methodologies, including participant demographics, dosage, treatment duration, and microbiome assessment techniques. This heterogeneity complicates direct comparisons and synthesis of results. Many studies on the gut microbiome are cross-sectional, providing a snapshot of microbiome composition at a single time point. These studies may fail to capture the dynamic changes in gut bacteria over the course of treatment, limiting insights into causality. Differences in microbiome sequencing technologies (e.g., 16S rRNA sequencing vs. shotgun metagenomics) and analytical pipelines could introduce variability in results, making it difficult to draw consistent conclusions. Many microbiome studies involve small cohorts, limiting statistical power and the ability to detect subtle or clinically meaningful changes in the gut microbiome. While the review discusses changes in microbiome composition, it lacks mechanistic insights and may not provide sufficient mechanistic understanding of how these changes interact with psychiatric treatments and contribute to treatment resistance or efficacy. While the review highlights immediate changes in gut bacteria associated with treatments, it may not sufficiently address long-term impacts, such as the potential development of antibiotic resistance or chronic dysbiosis.

### Gaps in current research

Potential sources of bias in these studies encompass variations in sample characteristics, notably the marked clinical heterogeneity within the BP cohort, geographical and dietary influences, and potential author overlap between papers (i.e., Flowers et al., – same groups have published most papers). As a result, it is challenging to interpret the disparities in measures of diversity and abundance between studies. Only four studies were interventional, where participants underwent a 3-4 week treatment with psychotropic medications (94,99,100,102). These studies employed a fixed dose of quetiapine (300mg/day) without a discernible dose-response correlation. The remaining eight studies were observational, where researchers monitored and compared the effects of psychotropic medications between individuals who were taking psychotropics and those who did not. There was significant heterogeneity between studies. Out of the total number of studies conducted (12 in total), only a subset shared comparability due to their utilization of a cross-sectional cohort design for comparing individuals with BP to HC groups. These studies generally featured modest sample sizes; in some studies, the patient group did not receive placebo treatment as a control (100).

Geographical variation may present another confounder since regional diets can vary greatly. While antibiotics, probiotics, and prebiotics were prohibited in many studies, it is essential to note that participants did not adhere to a standardized diet or physical activity, both pivotal environmental factors influencing gut microbiota composition (193). Some studies also allowed for a combination of medications in addition to quetiapine treatment, so underlying polypharmacy precludes us from drawing definitive conclusions. It would be difficult to control for this in future studies since most BP patients exist on an affective mood spectrum, which necessitates the use of multiple medications (144). Additionally, participant age was not matched between the patient and control groups and gut microbiome composition changes rapidly with age (145,146).

Only a subset of included studies was comparable; however, they often had small sample sizes, and in some cases, the patient group did not receive a placebo treatment as a control (100). None of the studies examined differences in AAP responses between BP-I and BP-II. Nevertheless, the findings may provide important preliminary information, as the study began at the time of diagnosis and continued with follow-up assessments throughout the treatment period. Some studies examined gut microbiota within the context of various psychiatric conditions, such as SCZ, alongside BP and explored diverse treatments, including starch and probiotics; however, the outcomes of these investigations fell beyond the scope of this review.

Another constraint pertained to the dropout rate among patients who initially received treatments, leading to the re-evaluation and final inclusion of only a subset of the patient population (96). Consequently, certain studies could not elucidate whether alterations in microbiota traits were tied to treatments. To establish a cause-and-effect relationship between BP gut microbiota and bipolar pharmacological treatments, a longitudinal cross-sectional cohort design is imperative. It is worth mentioning that the distinction between BP-I and BP-II in relation to gut microbiome composition changes following treatment was not examined in the studies.

### Future directions

The intestinal microbiome may govern varying drug responses between the sexes. An increasing number of microbiome studies have revealed endocrine bacteria that can produce and respond to hormones (e.g., 5-HT, DA, and estrogen) and regulate hormonal homeostasis by inhibiting gene transcription (e.g., prolactin) (147). Studies have also shown that females appear to have significantly higher levels of *Bifidobacteria* and *Bilophila* than males (148,149). This difference in baseline microbiome composition may explain why females under AAP treatment may express decreased gut microbiome species diversity (i.e., *Akkermansia muciniphila*) compared to HC, while males showed no difference (93). Some other studies have reported associations between the menstrual cycle and BP, particularly in a subgroup of females with enhanced hormonal sensitivity.

This population seems to experience heterogeneous menstrual cycle effects on depressive, hypomanic, and manic episodes (150), leading to greater chronobiological disruptions across the follicular and luteal phases (151). Sex-biased responses to psychotropics can also be due to inherent differences in physiology between sexes. Compared to females, on average, males have a higher dissolution rate, which is the rate-limiting step in drug absorption, as well as larger fluid volumes in the stomach and small intestine (following the normalization of body weight) (152). Females, on the other hand, have, on average, higher fasted state pH, lower acid secretion, smaller stomachs, and longer colonic transit time (153–155). These can all influence the ionization and solubility of pH-sensitive drugs, thereby affecting absorption and drug bioavailability (156,157). Notably, pre-menopausal females have a significantly longer gastric emptying time while post-menopausal females have a shorter time, which is more similar to average males (158).

As discussed previously and shown in the studies, many AAPs and antidepressants exhibit bactericidal and bacteriostatic activity and induce gut microbial antibiotic resistance (159–161). Although some antibiotics have been suggested as conjunctive therapies for BP, including tetracyclines (162), such as minocycline (163–165) and doxycycline (166–168), dangers from the global spread of antibiotic resistance should be considered. Plus, persistent use of psychotropics may also harm commensal beneficial gut bacteria. Accordingly, more caution should be made when prescribing psychotropics, which perhaps can be tapered off with alternative treatments such as cognitive behavioural therapies, education, and support groups (169,170).

It has not yet been possible to establish a healthy gut composition in individuals with BP, which can be defined by a balanced and diverse microbiome, strong intestinal integrity, and efficient digestive function; thus the interindividual gastrointestinal microbiome variations further complicate effective interventions (171,172). Dietary interventions may help enhance responses to drug therapy (173,174) and address metabolic side effects caused by the medications (75,78). Antioxidants and vitamin B supplements have been considered as adjuncts to antipsychotic drugs for SCZ patients (175), and high fibre intakes are correlated with reduced risk of depressive symptoms (176,177).

Furthermore, compared to non-vegetarians, vegetarians typically have a more rapid gastrointestinal transit time and gastric emptying (178). One of the most widely studied dietary interventions is the Mediterranean Diet, characterized by a high level of fibre, polyunsaturated fatty acids, anti-inflammatory and antioxidative products (179). These can increase *Faecalibacterium* strains (linked to butyrate metabolism), diminishing gut inflammation (180), reducing *Firmicutes* and *Proteobacteria phyla* (181), and stimulating fermentation in the lumen and adsorption of bile acid (178); therefore, this diet leads to better drug adsorption and bioavailability (81). Administering purified prebiotic Bimuno™ galacto-oligosaccharides powder with olanzapine can increase *Bifidobacterium* and attenuated weight gain induced by the AAP (75). Similarly, administering olanzapine with an antibiotic containing neomycin, metronidazole, and polymyxin B can attenuate weight gain and metabolic problems in rats (77).

Pro- and prebiotics have been used to reduce inflammation, decrease cardiovascular disease risks, and promote weight loss (182,183). They have also exhibited anxiolytic and antidepressant effects. In fact, a study of 80 first-episode drug-naive BP patients who underwent psychotropic therapy supplemented with probiotics showed clinical symptom improvements and reduced oxidative stress markers (allantoic acid, choline, creatine, hypoxanthine, inosine, uric acid) 3-month post-intervention (184). In another research report, probiotic supplementation improved attention, psychomotor processing speed, and executive functions also in 20 euthymic BP patients after 3 months (185).

Another innovative approach targeting gut microbiota in BP is faecal microbiota transplantation (FMT). This method involves transferring stool from a healthy donor into the pathological colon to restore a healthy gut microbiome and ameliorate systems (186). Studies on these in the psychiatry field are still limited (187). One significant success was reported in a case study where a BP patient, 6 months after undergoing FMT from their spouse, showed neither manic nor depressive symptoms (188). This method also entails some complicated challenges regarding donors and recipients’ compatibility as well as sources (189).

Lastly, there are some immune-based therapeutic strategies suggested for BP and MDD patients which do not follow the conventional treatment approach (190). For instance, celecoxib is a nonsteroidal anti-inflammatory drug commonly used to treat arthritis and joint pain but has been shown to be highly effective for bipolar depression in minimizing anxiety and facilitating treatment response (191). The same results were obtained with aspirin (192). N-acetylcysteine (NAC), a medication usually used to treat acetaminophen poisoning, was also proven to mitigate gut dysbiosis in mice (193) and improve depressive symptoms in BP patients (190,194).

## Conclusion

Medications such as antidepressants, antipsychotics, and anticholinergics significantly influence microbial physiology, altering the intestinal microbiome in ways that can impact therapeutic efficacy. These effects are particularly important to consider when developing drugs for neurological disorders, as microbial metabolism can either enhance or diminish treatment outcomes. Research on the psychotropic effects of medications on the gut microbiome in BP has produced mixed results, likely due to factors such as the anatomy and physiology of the gastrointestinal tract, the release and absorption profiles of the medications, polypharmacy, and the limitations of current study designs. Our systematic review of 12 studies highlights that psychotropic treatments, including lithium and quetiapine, promote the growth of beneficial gut bacteria associated with intestinal health but also increase the abundance of pathogenic bacteria linked to metabolic dysfunctions. These changes are more pronounced in female patients, who demonstrate greater microbial diversity shifts following treatment. Additionally, psychotropic treatments are associated with an increase in multidrug antibiotic resistance within the gut microbiome, raising concerns about potential long-term health risks.

Quetiapine responders—patients with improved depressive symptoms—exhibited distinct gut microbiome profiles that were more similar to those of healthy individuals compared to non-responders. This group also demonstrated neural connectivity patterns akin to healthy subjects, suggesting a possible gut-brain axis interaction contributing to treatment efficacy. Conversely, non-responders may possess inherent gut dysbiosis that compromises treatment outcomes. The role of *Clostridium bartlettii* in BP remains unclear, with mixed findings on its abundance following quetiapine treatment. These findings underscore the dual impact of psychotropic medications on gut microbiota, highlighting both potential therapeutic benefits and risks. The increased prevalence of beneficial bacteria alongside pathogenic and antibiotic-resistant strains reflects the complexity of microbial responses to treatment. While the mechanisms driving these shifts and their causal relationships remain uncertain, this analysis offers promising directions for improving treatment outcomes in BP. Leveraging insights into gut microbiome dynamics could lead to more personalized and effective therapies, reducing side effects and addressing the diverse needs of BP patients.

## Data Availability

All data produced in the present work are contained in the manuscript

## List of figures and tables

**S1 Table.** Search strategies by database.

**S2 Table.** Certainty of evidence assessment.

**S3 Table.** PRISMA 2020 abstract checklist.

**S4 Table.** PRISMA 2020 checklist.

## Author statements

### Author contributions

Truong An Bui: Conceptualization, Formal analysis, Validation, Data curation, Visualization Writing; Benjamin R. O’Croinin: Formal analysis, Validation, Data Curation, Writing – Review and Editing; Liz Dennett: Methodology, Software, Writing – Review and Editing; Ian R. Winship: Writing – Review and Editing, Supervision; Andrew Greenshaw: Conceptualization, Writing – Review and Editing, Supervision.

### Availability of data

The data supporting the findings of this study are available within the article and its supplementary materials.

