## Supplementary material for "Pharmaco-psychiatry and gut microbiome: A systematic review of effects of psychotropic drugs for bipolar disorder": S1 Table

**S1 Table.** Database search queries.

| Date | Database | Return |
|---|---|---|
| 1946 to 2024 August 13 | **Ovid MEDLINE(R) ALL** | |
| | 1 "bipolar and related disorders"/ or bipolar disorder/ | 46779 |
| | 2 (bipolar or manic depress* or mania).mp. | 99622 |
| | 3 1 or 2 | 99622 |
| | 4 gastrointestinal microbiome/ or ((gastrointestinal or intestin* or gut or bowel* or digesti* or colon or colonic or feces or faeces or fecal or faecal) and (flora or microb* or microflora or metagenom* or bacteri* or coloniz* or colonis*)).mp. | 304593 |
| | 5 metagenome/ or exp bacteria/ or exp microbiota/ | 1644121 |
| | 6 gastrointestinal tract/ or exp intestines/ or exp lower gastrointestinal tract/ | 454703 |
| | 7 4 or (5 and 6) | 310797 |
| | 8 antimanic agents/ or carbamazepine/ or gabapentin/ or lithium carbonate/ or lithium chloride/ or lithium compounds/ or valproic acid / or antipsychotic agents/ | 99118 |
| | 9 (Mood stabilizer* or mood stabiliser* or antipsychotic* or anti-psychotic* or anti-manic or antimanic or haloperidol or haldol or loxapine or loxitane or loxapine or adasuve or Aripiprazole or Abilify or Asenapine or Saphris or Cariprazine or Vraylar or Lumateperone or Caplyta or Lurasidone or Latuda or Olanzapine or Zyprexa or Olanzapine or samidorphan or Lybalvi or Quetiapine or Seroquel or risperidone or Risperdal or Ziprasidone or Geodon or Lithium or Divalproex or Valproic acid or Valproate or Carbamazepine or Oxcarbazepine or Lamotrigine or Fluoxetine or Symbyax or gabapentin).mp. | 237786 |
| | 10 8 or 9 | 237786 |
| | 11 3 and 7 and 10 | 26 |
| 1806 to April Week 2 2024 | **APA PsycInfo** | |
| | 1 exp bipolar disorder/ | 36298 |
| | 2 (bipolar or manic depress* or mania).mp. | 63150 |
| | 3 1 or 2 | 63343 |
| | 4 (gastrointestinal or intestin* or gut or bowel* or digesti* or colon or colonic or feces or faeces or fecal or faecal) and (flora or microb* or microflora or metagenom* or bacteri* or coloniz* or colonis*)).mp. | 3465 |
| | 5 exp Valproic Acid / or exp Mood Stabilizers/ or exp Carbamazepine/ or exp Lithium/ | 10783 |
| | 6 (Mood stabilizer* or mood stabiliser* or antipsychotic* or anti-psychotic* or anti-manic or antimanic or haloperidol or haldol or loxapine or loxitane or loxapine or adasuve or Aripiprazole or Abilify or Asenapine or Saphris or Cariprazine or Vraylar or Lumateperone or Caplyta or Lurasidone or Latuda or Olanzapine or Zyprexa or Olanzapine or samidorphan or Lybalvi or Quetiapine or Seroquel or risperidone or Risperdal or Ziprasidone or Geodon or Lithium or Divalproex or Valproic acid or Valproate or Carbamazepine or Oxcarbazepine or Lamotrigine or Fluoxetine or Symbyax or gabapentin).mp. | 81284 |
| | 7 5 or 6 | 81284 |
| | 8 3 and 4 and 7 | 12 |
| | **SCOPUS (Advanced search)** | |
| | TITLE-ABS-KEY(bipolar or manic-depress* or mania) AND TITLE-ABS-KEY((gastrointestinal or intestin* or gut or bowel* or digesti* or colon or colonic or feces or faeces or fecal or faecal) and (flora or microb* or microflora or metagenom* or bacteri* or coloniz* or colonis*)) AND TITLE-ABS-KEY(Mood-stabilizer* or mood-stabiliser* or antipsychotic* or anti-psychotic* or anti-manic or antimanic or haloperidol or haldol or loxapine or loxitane or loxapine or adasuve or Aripiprazole or Abilify or Asenapine or Saphris or Cariprazine or Vraylar or Lumateperone or Caplyta or Lurasidone or Latuda or Olanzapine or Zyprexa or Olanzapine or samidorphan or Lybalvi or Quetiapine or Seroquel or risperidone or Risperdal or Ziprasidone or Geodon or Lithium or Divalproex or Valproic-acid or Valproate or Carbamazepine or Oxcarbazepine or Lamotrigine or Fluoxetine or Symbyax or gabapentin) | 123 |
| 1974 to 2024 August 13 | **Embase** | |
| | 1 exp bipolar disorder/ | 86071 |
| | 2 (bipolar or manic depress* or mania).mp. | 161200 |
| | 3 1 or2 | 161512 |
| | 4 exp intestine flora/ | 120047 |
| | 5 microflora/ or feces microflora/ | 52160 |
| | 6 exp intestine/ | 542783 |
| | 7 exp gastrointestinal tract/ | 82052 |
| | 8 5 and (6 or 7) | 9469 |
| | 9 (gastrointestinal or intestin* or gut or bowel* or digesti* or colon or colonic or feces or faeces or fecal or faecal) and (flora or microb* or microflora or metagenom* or bacteri* or coloniz* or colonis*)).mp. | 379936 |
| | 10 4 or 8 or 9 | 380074 |
| | 11 exp mood stabilizer/ | 150399 |
| | 12 (Mood stabilizer* or mood stabiliser* or antipsychotic* or anti-psychotic* or anti-manic or antimanic or haloperidol or haldol or loxapine or loxitane or loxapine or adasuve or Aripiprazole or Abilify or Asenapine or Saphris or Cariprazine or Vraylar or | 457304 |

| | | |
|---|---|---|
| | Lumateperone or Caplyta or Lurasidone or Latuda or Olanzapine or Zyprexa or Olanzapine or samidorphan or Lybalvi or Quetiapine or Seroquel or risperidone or Risperdal or Ziprasidone or Geodon or Lithium or Divalproex or Valproic acid or Valproate or Carbamazepine or Oxcarbazepine or Lamotrigine or Fluoxetine or Symbyax or gabapentin).mp.<br>13 11 or 12<br>14 3 and 10 and 13 | <br><br><br><br><br>457490<br>137 |
| 13 August 2024 | **PubMed** | |
| | Search: ((bipolar[Title/Abstract] OR manic-depress*[Title/Abstract] OR mania[Title/Abstract]) AND (gastrointestinal[Title/Abstract] OR intestin*[Title/Abstract] OR gut[Title/Abstract] OR bowel*[Title/Abstract] OR digesti*[Title/Abstract] OR colon[Title/Abstract] OR colonic[Title/Abstract] OR feces[Title/Abstract] OR faeces[Title/Abstract] OR fecal[Title/Abstract] OR faecal[Title/Abstract])) AND (Mood-stabilizer*[Title/Abstract] OR mood-stabiliser*[Title/Abstract] OR antipsychotic*[Title/Abstract] OR anti-psychotic*[Title/Abstract] OR anti-manic[Title/Abstract] OR antimanic[Title/Abstract] OR haloperidol[Title/Abstract] OR haldol[Title/Abstract] OR loxapine[Title/Abstract] OR loxitane[Title/Abstract] OR loxapine[Title/Abstract] OR adasuve[Title/Abstract] OR Aripiprazole[Title/Abstract] OR Abilify[Title/Abstract] OR Asenapine[Title/Abstract] OR Saphris[Title/Abstract] OR Cariprazine[Title/Abstract] OR Vraylar[Title/Abstract] OR Lumateperone[Title/Abstract] OR Caplyta[Title/Abstract] OR Lurasidone[Title/Abstract] OR Latuda[Title/Abstract] OR Olanzapine[Title/Abstract] OR Zyprexa[Title/Abstract] OR Olanzapine[Title/Abstract] OR samidorphan[Title/Abstract] OR Lybalvi[Title/Abstract] OR Quetiapine[Title/Abstract] OR Seroquel[Title/Abstract] OR risperidone[Title/Abstract] OR Risperdal[Title/Abstract] OR Ziprasidone[Title/Abstract] OR Geodon[Title/Abstract] OR Lithium[Title/Abstract] OR Divalproex[Title/Abstract] OR Valproic-acid[Title/Abstract] OR Valproate[Title/Abstract] OR Carbamazepine[Title/Abstract] OR Oxcarbazepine[Title/Abstract] OR Lamotrigine[Title/Abstract] OR Fluoxetine[Title/Abstract] OR Symbyax[Title/Abstract] OR gabapentin[Title/Abstract]) | 171 |
