## Supplementary material for "Pharmaco-psychiatry and gut microbiome: A systematic review of effects of psychotropic drugs for bipolar disorder": S2 Table

**Psychotropics compared to non-psychotropics for bipolar disorder: Correlation between microbial species and psychotropics**

**Patient or population:** bipolar disorder
**Setting:** Inpatient, outpatient, and community
**Intervention:** Psychotropics
**Comparison:** Non-psychotropics

| Outcomes | № of participants (studies) | Study design | Intervention (n) | Certainty of the evidence (GRADE) | Findings |
|---|---|---|---|---|---|
| **Olanzapine v. Klebsiella** (*Ioannou et al., 2024*)<br>assessed with: Metagenomic shotgun sequencing | 103<br>(1 non-randomised study) | cross-sectional | 2.4 | ⊕⊕⊕◯<br>Moderate* | Positive correlation<br>beta = 0.07, SE = 0.02, p = 0.01 |
| **Lithium v. Anaeromassilibacillus** (*Ioannou et al., 2024*)<br>assessed with: Metagenomic shotgun sequencing | 103<br>(1 non-randomised study) | cross-sectional | 38 | ⊕⊕⊕⊕<br>High | Positive correlation<br>beta = 1.28, SE = 0.30, p = 0.005 |
| **Atypical antipsychotics v. Lachnospiraceae** (*Flowers et al., 2017*)<br>assessed with: 16S ribosomal RNA sequencing | 117<br>(1 non-randomised study) | cross-sectional | 49 | ⊕⊕⊕◯<br>Low** | Positive correlation<br>p = 0.001 |
| **Atypical antipsychotics v. Akkermansia** (*Flowers et al., 2017*)<br>assessed with: 16S ribosomal RNA sequencing | 117<br>(1 non-randomised study) | cross-sectional | 49 | ⊕⊕⊕◯<br>Low** | Negative correlation<br>p = 0.03 |
| **Quetiapine v. Clostridium bartlettii** (*Xi et al., 2023*)<br>assessed with: Metagenomic sequencing<br>follow-up: 4 weeks | 75<br>(1 non-randomised study) | before-after | 43 | ⊕⊕◯◯<br>Low*** | Negative correlation<br>Statistics not reported |
| **Quetiapine v. Bacteroides sp_2_1_22** (*Xi et al., 2023*)<br>assessed with: Metagenomic sequencing<br>follow-up: mean 4 weeks | 75<br>(1 non-randomised study) | before-after | 43 | ⊕⊕◯◯<br>Low*** | Negative correlation<br>Statistics not reported |
| **Quetiapine v. Bacteroides sp_3_1_19** (*Xi et al., 2023*)<br>assessed with: Metagenomic sequencing<br>follow-up: 4 weeks | 75<br>(1 non-randomised study) | before-after | 43 | ⊕⊕◯◯<br>Low*** | Negative correlation<br>Statistics not reported |
| **Quetiapine v. Eubacterium biforme** (*Xi et al., 2023*)<br>assessed with: Metagenomic sequencing<br>follow-up: 4 weeks | 75<br>(1 non-randomised study) | before-after | 43 | ⊕⊕◯◯<br>Low*** | Positive correlation<br>Statistics not reported |
| **Quetiapine v. Weissella confusa** (*Xi et al., 2023*)<br>assessed with: Metagenomic sequencing<br>follow-up: 4 weeks | 75<br>(1 non-randomised study) | before-after | 43 | ⊕⊕◯◯<br>Low*** | Positive correlation<br>Statistics not reported |
| **Quetiapine v. Oribacterium sinus** (*Xi et al., 2023*)<br>assessed with: Metagenomic sequencing<br>follow-up: 4 weeks | 75<br>(1 non-randomised study) | before-after | 43 | ⊕⊕◯◯<br>Low*** | Positive correlation<br>Statistics not reported |
| **Quetiapine v. Barnesiella intestinihominis** (*Xi et al., 2023*)<br>assessed with: Metagenomic sequencing<br>follow-up: 4 weeks | 75<br>(1 non-randomised study) | before-after | 43 | ⊕⊕◯◯<br>Low*** | Positive correlation<br>Statistics not reported |
| **Quetiapine v. Bifidobacterium dentium** (*Lai et al., 2022*)<br>assessed with: Metagenomic sequencing<br>follow-up: 4 weeks | 62<br>(1 non-randomised study) | before-after | 62 | ⊕⊕◯◯<br>Low*** | Negative correlation<br>Statistics not reported |
| **Quetiapine v. Enterococcus hirae** (*Lai et al., 2022*)<br>assessed with: Metagenomic sequencing<br>follow-up: 4 weeks | 62<br>(1 non-randomised study) | before-after | 62 | ⊕⊕◯◯<br>Low*** | Negative correlation<br>Statistics not reported |
| **Quetiapine v. Anaerofustis stercorihominis** (*Lai et al., 2022*)<br>assessed with: Metagenomic sequencing<br>follow-up: 4 weeks | 62<br>(1 non-randomised study) | before-after | 62 | ⊕⊕◯◯<br>Low*** | Positive correlation<br>Statistics not reported |

| | | | | | |
|---|---|---|---|---|---|
| **Quetiapine v. Streptococus cristatus** (*Lai et al., 2022*)<br>assessed with: Metagenomic sequencing<br>follow-up: 4 weeks | 62<br>(1 non-randomised study) | before-after | 62 | ⊕⊕◯◯<br>Low*** | Positive correlation<br>Statistics not reported |
| **Quetiapine v. Campylobacter hominis** (*Lai et al., 2022*)<br>assessed with: Metagenomic sequencing<br>follow-up: 4 weeks | 62<br>(1 non-randomised study) | before-after | 62 | ⊕⊕◯◯<br>Low*** | Positive correlation<br>Statistics not reported |
| **Quetiapine v. Porphyromonas uenonis** (*Lai et al., 2022*)<br>assessed with: Metagenomic sequencing<br>follow-up: 4 weeks | 62<br>(1 non-randomised study) | before-after | 62 | ⊕⊕◯◯<br>Low*** | Positive correlation<br>Statistics not reported |
| **Quetiapine v. Anaerococcus vaginalis** (*Lai et al., 2022*)<br>assessed with: Metagenomic sequencing<br>follow-up: 4 weeks | 62<br>(1 non-randomised study) | before-after | 62 | ⊕⊕◯◯<br>Low*** | Positive correlation<br>Statistics not reported |
| **Quetiapine v. Clostridium perfringens** (*Lai et al., 2022*)<br>assessed with: Metagenomic sequencing<br>follow-up: 4 weeks | 62<br>(1 non-randomised study) | before-after | 62 | ⊕⊕◯◯<br>Low*** | Positive correlation<br>Statistics not reported |
| **Quetiapine v. Streptococcus parasanguinis** (*Lai et al., 2022*)<br>assessed with: Metagenomic sequencing<br>follow-up: 4 weeks | 62<br>(1 non-randomised study) | before-after | 62 | ⊕⊕◯◯<br>Low*** | Positive correlation<br>Statistics not reported |
| **Quetiapine v. Costridium scindens** (*Lai et al., 2022*)<br>assessed with: Metagenomic sequencing<br>follow-up: 4 weeks | 62<br>(1 non-randomised study) | before-after | 62 | ⊕⊕◯◯<br>Low*** | Positive correlation<br>Statistics not reported |
| **Quetiapine v. Streptococcus salivarius** (*Lai et al., 2022*)<br>assessed with: Metagenomic sequencing<br>follow-up: 4 weeks | 62<br>(1 non-randomised study) | before-after | 62 | ⊕⊕◯◯<br>Low*** | Positive correlation<br>Statistics not reported |
| **Quetiapine v. Ruminococcus obeum** (*Lai et al., 2022*)<br>assessed with: Metagenomic sequencing<br>follow-up: 4 weeks | 62<br>(1 non-randomised study) | before-after | 62 | ⊕⊕◯◯<br>Low*** | Positive correlation<br>Statistics not reported |
| **Quetiapine v. Enterobacter cloacae** (*Lai et al., 2022*)<br>assessed with: Metagenomic sequencing<br>follow-up: 4 weeks | 62<br>(1 non-randomised study) | before-after | 62 | ⊕⊕◯◯<br>Low*** | Positive correlation<br>Statistics not reported |
| **Quetiapine v. Eubacterium rectale** (*Lu et al.2019*)<br>assessed with: qPCR<br>follow-up: 4 weeks | 36<br>(1 non-randomised study) | before-after | 36 | ⊕⊕⊕⊕<br>High | Positive correlation<br>p = 0.004 |

*Serious imprecision. 2.4 olanzapine equivalent users reported for 103 participants do not meet the optimal information size as calculated.
**Some concerns of risk of bias due to missing statistic details, some uncontrolled confounding factors, and inconsistency in data reporting.
***Some inconsistency in follow-up measurement. High risk of bias due to lack of statistics and uncontrolled confounding factors.

**GRADE Working Group grades of evidence**
**High certainty:** we are very confident that the true effect lies close to that of the estimate of the effect.
**Moderate certainty:** we are moderately confident in the effect estimate: the true effect is likely to be close to the estimate of the effect, but there is a possibility that it is substantially different.
**Low certainty:** our confidence in the effect estimate is limited: the true effect may be substantially different from the estimate of the effect.
**Very low certainty:** we have very little confidence in the effect estimate: the true effect is likely to be substantially different from the estimate of effect.
